# The changing landscape of respiratory viruses contributing to respiratory hospitalisations: results from a hospital-based surveillance in Quebec, Canada, 2012-13 to 2021-22

**DOI:** 10.1101/2022.07.01.22277061

**Authors:** Rodica Gilca, Rachid Amini, Sara Carazo, Charles Frenette, Guy Boivin, Hugues Charest, Jeannot Dumaresq

**Affiliations:** Department of biological risks, Institut national de sante publique du Québec, Québec City, Québec, Canada; Research Center of CHU de Québec-Université Laval, Québec City, Québec, Canada; Department of Medicine, Laval University, Québec City, Québec, Canada; Department of Medicine, Division of Infectious Diseases, McGill University Health Center, McGill University, Montreal, Canada; Laboratoire de santé publique du Québec, Institut national de sante publique du Québec, Montréal, Québec, Canada; Departement of Microbiology and Infectiology, CISSS de Chaudière-Appalaches, Lévis, Québec, Canada

**Keywords:** Respiratory viruses, SARS-CoV-2, hospitalisations, acute respiratory infections, children, adults, coinfections, pre-pandemic, pandemic

## Abstract

**Background:** A comprehensive description of the combined effect of SARS-CoV-2 and respiratory viruses (RV) other than SARS-CoV-2 (ORV) on hospitalisations is lacking.

**Aim:** To compare viral etiology of acute respiratory infections (ARI) hospitalisations before and during two pandemic years from a surveillance network in Quebec, Canada.

**Method:** We compared detection of ORV and SARS-CoV-2 during 2020-21 and 2021-22 to 8 pre-pandemic influenza seasons in patients hospitalised with ARI who were tested systematically by a multiplex PCR.

**Results:** During pre-pandemic influenza seasons, overall RV detection was 92.7% (1,493) (48.3% respiratory syncytial virus (RSV)) in children and 62.8% (4,339) (40.1% influenza) in adults. Overall RV detection in 2020-21 was 58.6% (29) in children (all ORV) and 43.7% (333) in adults (3.4% ORV, 40.3% SARS-CoV2, both including coinfections). In 2021-22 overall RV detection was 91.0% (201) in children (82.8% ORV, 8.1% SARS-CoV-2, both including coinfections) and 55.5% (527) in adults (14.1% ORV, 41.4% SARS-CoV-2, both including coinfections).

Virtually no influenza was detected in 2020-21 and in 2021-22 up to epi-week 2022-9 presented here; no RSV was detected in 2020-21. In 2021-22, detection of RSV was comparable to pre-pandemic years but with an unusually early season. There were significant differences in ORV and SARS-CoV-2 detection between time periods and age groups.

**Conclusion:** Significant continuous shifts in age distribution and viral etiology of ARI hospitalisations occurred during two pandemic years. This reflects evolving RV epidemiology and underscores the need for increased scrutiny of ARI hospitalisation etiology to inform tailored public health recommendations.

## Introduction

Stringent mitigation efforts such as border closures and travel restrictions, lockdowns, social distancing, use of masks in public spaces, school and business closures, and teleworking have been implemented worldwide to reduce the transmission of severe acute respiratory syndrome virus 2 (SARS-CoV-2) and its impact on hospital bed capacity. While first weeks of 2020 in the Northern hemisphere were dominated by respiratory viruses other than SARS-CoV-2 (ORV), SARS-CoV-2 almost completely replaced seasonally circulating ORV within several weeks (1–3), with a subsequent alteration of traditional seasonality of some of them and virtual disappearance of others during extended periods of time in different parts of the world (4–9). One of the collateral consequences observed early during the pandemic was the decrease in pediatric visits and hospitalizations overall and especially of those associated with acute respiratory infections (ARI)(10,11), bronchiolitis(12,13), as well as pediatric asthma exacerbations associated with ARI(14).

The pandemic had an impact on resources with decreased volumes of performed tests and changes in the propensity to test for ORV, as well as on the health-seeking behavior, which may complicate the interpretation of ORV surveillance. Detection of ORV in hospitalised patients may be more informative since admission requires a certain degree of severity and propensity to be tested for a larger panel of respiratory viruses is higher. However, because of high demand of SARS-CoV-2 tests on hospital laboratories, the testing for ORV has been reduced even in hospitalised patients during the pandemic. A number of reports described detection of ORV in patients hospitalised with ARI or with COVID-19 during the pandemic(8,15–19). However, to our knowledge none described results of systematic detection of both ORV and SARS-CoV-2 (not only at physician request) using a panel of multiple RV in a multicenter network including both pandemic years and comparing them with as long as 8 pre-pandemic years. The characterisation of combined impact of both SARS-CoV-2 and ORV on hospital capacity during the two pandemic years and its comparison with pre-pandemic seasons may provide insightful information on post-pandemic period when the SARS-CoV-2 will cocirculate along with ORV.

In Quebec, Canada, a prospective hospital-based surveillance network with systematic testing for a panel of 17 respiratory viruses in pediatric and adult patients admitted for ARI has been in place since 2012-2013 during periods with high influenza circulation (20–23). The same network was used for the surveillance during the pandemic, by adding the SARS-CoV-2 to the panel. We report here the results for two pandemic winter seasons (2020-21 and 2021-22) and compare them to eight previous seasons.

## Method

### Study Population

The study design during the pre-pandemic years has been described in detail elsewhere(20–23). In brief, four regional hospitals (2 community, 2 academic/tertiary; all of them serving both children and adults) with a catchment area of nearly 10% of the Quebec population (approximately 8.6 million in 2021) participated in the surveillance during eight influenza seasons since 2012-13. In 2020-21, one of the 4 hospitals was not able to participate (≈15% of the population included in previous years) because of the challenges with hospital resources during the pandemic; this hospital rejoined the network in 2021-22. Two additional tertiary hospitals, one adult and one pediatric, joined the network in 2021-22, for a total of 6 hospitals (≈15% of the Quebec population). Results from these two hospitals are included only in the description of virus detection per week and are not used for the comparison with pre-pandemic period. All patients presenting to their emergency department with ARI were systematically swabbed during high influenza activity weeks of pre-pandemic years or during periods with increasing hospitalisations due to SARS-CoV-2 during the pandemic. Eligible patients were those admitted for ≥24 hours with a standardized definition (fever/feverishness not attributed to other illness or cough or sore throat), expanded in 2020-21 to include symptoms specific for COVID-19 (fever/history of fever not attributed to other illness, or cough (or exacerbation of cough)/difficulty breathing (or exacerbation of difficulty breathing), or sudden extreme fatigue, or at least two of the following symptoms: rhinorrhea/nasal congestion, sore throat, myalgia/arthralgia, or sudden anosmia/ageusia). Nurses collected demographic and clinical details from the patient or legal representative on a standardized questionnaire and reviewed patients’ charts at discharge for additional clinical information.

### Surveillance period

For the pre-pandemic years, the surveillance period started when the positivity rate for influenza in respiratory specimens from the provincial sentinel laboratory surveillance was ≥15% for two consecutive weeks and stopped the week after this rate dropped below 15% or when the planned sample size for the season was achieved (800-1000 specimens depending on the season). The provincial laboratory surveillance included >40 laboratories across the province of Quebec with >100,000 respiratory specimens per year. Surveillance lasted from 7 to 12 weeks per season (median of 8,5 weeks), between epi-weeks 49 (earliest) and 14 (latest) (**Supplementary Figure 1**). In 2020-21, the surveillance started on September 27, 2020 (epi-week 40), during the ascending phase of the 2^nd^ COVID-19 wave in Québec and was halted on May 29, 2021 (epi-week 21) (overall duration 35 weeks) (**Figure 1, Supplementary Figure 1, Supplementary Table 1**). This period included circulation of ancestral and alpha SARS-CoV-2 variants(24). In 2021-22, the surveillance started on October 04, 2021 (epi-week 40), during the ascending phase of unusual interseason RSV surge in Québec; it included the descending phase of the 4^th^ wave (due to Delta variant); the 5^th^ Omicron wave and the ascending phase of the 6^th^ wave (due to BA2 variant); and it was halted on June 18 (epi-week 24). The 2 additional hospitals joined the surveillance starting at week 43 (adult hospital) and week 45 (pediatric hospital). Data reported here are those until March 05, 2022 (epi-week 9) (overall duration 22 weeks) and do not include unusually late influenza season in Quebec which started on epi-week 12.

**Figure 1.**
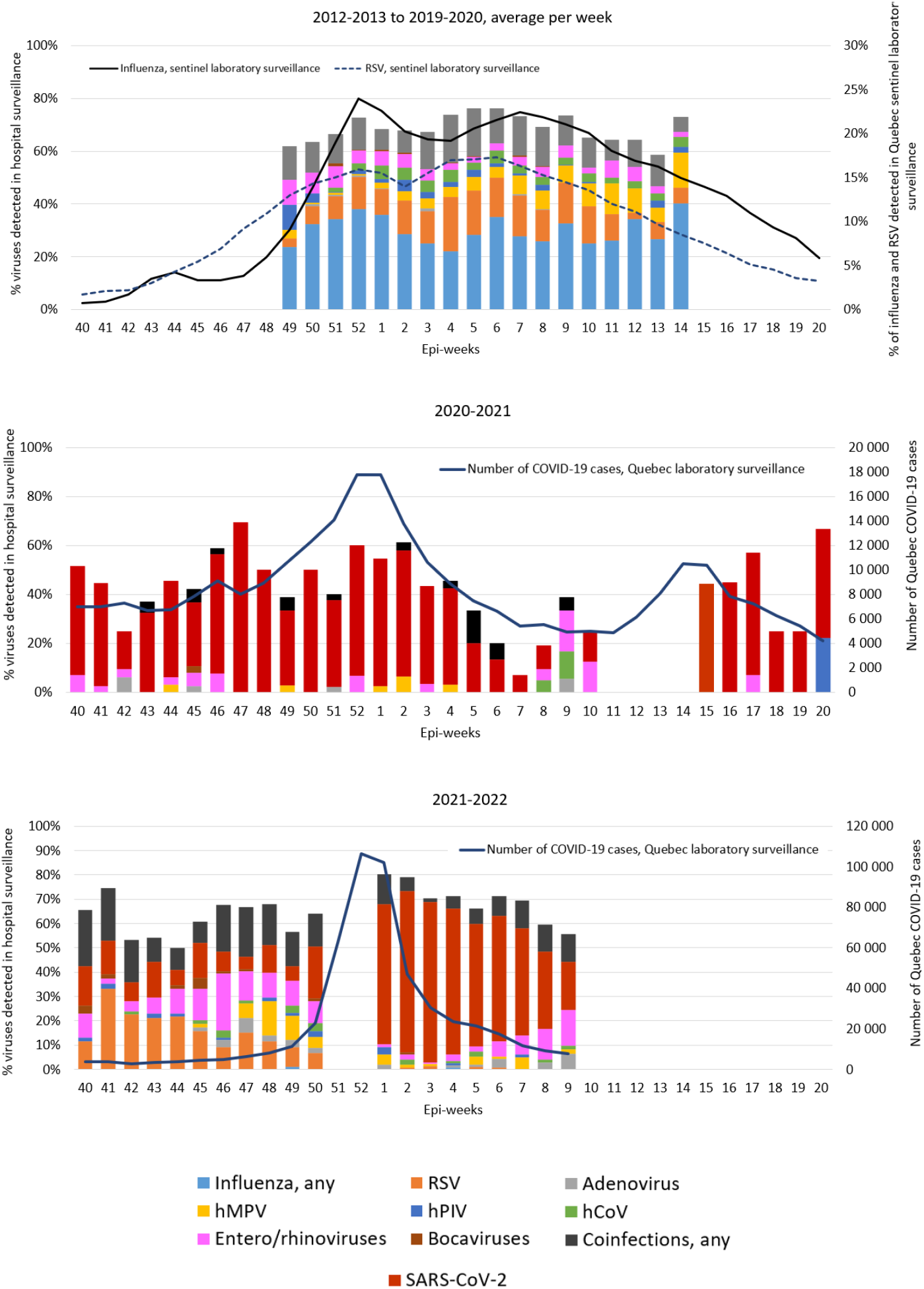
Proportions of respiratory virus detection in patients hospitalised for acute respiratory infections by epi-week in Quebec, Canada (epi-weeks 40 to 20 are presented) Abbreviations: hMPV: human metapneumovirus; RSV: respiratory syncytial virus; hPIV: human parainfluenza viruses; hCoV: common human coronaviruses Note to Figure 1: All participating hospitals are included (4 hospitals in 2012-13 to 2019-20 and 2020-2021; 6 hospitals in 2021-2022). In order to simplify the presentation, epi-weeks 53 (2014-15 and 2019-20) are excluded. For the purpose of the figure, weeks 40 to 20 are presented. Details on number of participants are presented in **Supplementary Figure 1**

Because of challenges with hospital resources during periods of high SARS-CoV-2 circulation, the surveillance was paused during some weeks and sampling of enrolment during pre-determined days of week was adopted by some hospitals (**Supplementary Figure 1**).

### Laboratory Analysis

Nasal specimens collected on flocked swabs from eligible patients were sent to the provincial public health laboratory (Laboratoire de Santé Publique du Québec, LSPQ) and tested using the Luminex® NxTAG Respiratory Pathogen Panel assay which detects influenza A (subtypes H3 and H1), influenza B, respiratory syncytial virus (RSV) A and B, human parainfluenza viruses (hPIV) 1, 2, 3, and 4, human metapneumovirus (hMPV), common human coronaviruses (HCoV) NL63, HKU1, 229E, and OC43, enteroviruses/rhinoviruses (not differentiated), adenovirus, bocavirus, and 3 bacteria (*Mycoplasma pneumoniae, Chlamydia pneumoniae*, and *Legionella pneumophila*). This assay was systematically used during all pre-pandemic years, starting from week 45 in 2020-21 and during the entire 2021-22 season. BioFire® Respiratory Panel 2.1 (RP2.1) was used for ORV testing by local laboratories between epi-week 40 and 44 in 2020. In-house multiplex reverse transcription PCR (MRVP) detecting influenza A and B, hPIV 1-2-3, adenovirus, rhinovirus, enterovirus, hCoV 229E and OC43, RSV, and hMPV was used by the adult center(25). In house PCR using LightMix® Modular Assays according to the manufacturer’s recommendations(26) was used by the pediatric center to detect influenza A and B, RSV, hCoV (not differentiated), hMPV, adenovirus, hPIV (not differentiated), enteroviruses/rhinoviruses (not differentiated), and SARS-CoV-2.

### Statistical Analysis

Proportions were compared by using chi-square or Fisher exact test when appropriate. Statistical significance was set at p<0.05. Statistical analyses were conducted using SAS, version 9.4 (SAS Institute, Cary, NC). Similar hospitalisation rate and viral etiology distribution was assumed for days with and without enrolment during weeks with only three enrolment days.

### Ethics

Institutional Review Board approval was obtained from all participating hospitals (Hôpital régional de Rimouski, Hôpital de Chicoutimi, Hôpital de la Cité-de-la-Santé – Laval, Centre hospitalier universitaire régional de Trois-Rivières) for the first 3 years and a signed informed consent was used. A waiver was obtained for the following years when the project was conducted as a sentinel surveillance mandated by the Ministry of Health from the Research Ethics Board of the Centre hospitalier universitaire de Québec-Université Laval.

## Results

### Participants to the surveillance

Overall, 9,339 patients potentially eligible for the surveillance were approached (6,412 during pre-pandemic period, 1,454 in 2020-21, and 1,473 in 2021-22) (see details in **Supplementary Figure 2**). A total of 7,793 patients hospitalised for community-acquired ARI were included in the analysis: 5,832 (1,493 children 0-17 years, 4,339 adults) during the 8 pre-pandemic influenza seasons, 791 (29 children, 762 adults) during the 2020-21 season and 1,170 (221 children, 949 adults) during the 2021-22 season (**Table 1**). In 2020-21, significantly less children were hospitalised with ARI (4% of all hospitalised patients) compared to previous seasons (26%, p<0,0001). During both pre-pandemic and pandemic winter seasons, very few young adults were hospitalised with ARI, with approximately 1% of 18-29-year-olds and 2% of 30-39-year-olds and a subsequent gradual increase with age (**Figure 2**). Compared to the pre-pandemic period, during the 1^st^ pandemic year (2020-21) the proportion of 18-59-year-olds was similar (14% vs 13% in pre-pandemic period, p>0.05), but it was higher for 60-79-year-olds (43% vs 32%, p<0.0001) and for ≥80-year-olds (39% vs 29%, p<0.0001). During the 2^nd^ pandemic year (2021-22), these proportions were lower or comparable to those from pre-pandemic period for children, 18-59-year-olds, and ≥80-year-olds (respectively 19% vs 26% (p<0.0001), 10% vs 13% (<0.001), and 30% vs 29%, p>0.05) but were higher for 60-79-year-olds (41% vs 32%, p<0.0001) (**Table 1, Figure 2**).

**Table 1.** Number and proportion of patients hospitalised for acute respiratory infections by age group and detected respiratory virus in Quebec, Canada, pre-pandemic and pandemic winter seasons

| Age group, years | Peaks of 2012-20 influenza seasons<br>(pre-pandemic, 4 hospitals) |  | 2020-21<br>(pandemic, 3 hospitals) |  |  |  | 2021-22<br>(pandemic, 4 hospitals) |  |  |  |
| --- | --- | --- | --- | --- | --- | --- | --- | --- | --- | --- |
|  | Number tested, n(%) | At least one virus positive, n(%) | Number tested, n(%) | At least one virus positive, n(%) | SARS-CoV-2**, n(%) | Other virus without SARS-CoV-2, n (%) | Number tested, n(%) | At least one virus positive, n(%) | SARS-CoV-2**, n(%) | Other virus without SARS-CoV-2, n (%) |
| 0-17 | 1493 (26%) | 1384 (93%) | 29 (4%) | 17 (**59%<sup>a b</sup>**) | 0 (0%) | 17 (59%) | 221 (19%) | 201 (91%) | 18 (8%) | 183 (83%) |
| 18-29 | 79 (1%) | 50 (63%) | 12 (2%) | 3 (**25%<sup>a</sup>**) | 2 (17%) | 1 (8%) | 12 (1%) | 7 (58%) | 6 (50%) | 1 (8%) |
| 30-39 | 130 (2%) | 75 (58%) | 13 (2%) | 6 (46%) | 6 (46%) | 0 (0%) | 17 (1%) | 13 (76%) | 10 (59%) | 3 (18%) |
| 40-49 | 160 (3%) | 104 (65%) | 29 (4%) | 16 (55%) | 16 (55%) | 0 (0%) | 20 (2%) | 14 (70%) | 14 (70%) | 0 (0%) |
| 50-59 | 409 (7%) | 267 (65%) | 55 (7%) | 28 (51%) | 26 (47%) | 2 (4%) | 66 (6%) | 39 (59%) | 31 (47%) | 8 (12%) |
| 60-69 | 744 (13%) | 452 (61%) | 147 (19%) | 48 (**33%<sup>ab</sup>**) | 44 (30%) | 4 (3%) | 197 (17%) | 92 (**47%<sup>a</sup>**) | 69 (35%) | 23 (12%) |
| 70-79 | 1111 (19%) | 678 (61%) | 194 (25%) | 78 (**40%<sup>ab</sup>**) | 73 (38%) | 5 (3%) | 286 (24%) | 146 (**51%<sup>a</sup>**) | 102 (36%) | 44 (15%) |
| ≥80 | 1706 (29%) | 1097 (64%) | 312 (39%) | 154 (**49%<sup>ab</sup>**) | 140 (45%) | 14 (4%) | 351 (30%) | 216 (62%) | 161 (46%) | 55 (16%) |
| Total | 5832 (100%) | 4107 (70%) | 791 (100%) | 350 (**44%<sup>ab</sup>**) | 307 (39%) | 43 (5%) | 1170 (100%) | 728 (**62%<sup>a</sup>**) | 411 (35%) | 317 (27%) |

\*Peaks of influenza seasons; \*\*with or without ORV; **In bold:** statistically significant difference;
<sup>a</sup> p value <0.05 for comparison with 2012-20, Fisher exact test;
<sup>b</sup> p value <0.05 for comparison between 2020-21 and 2021-22, Fisher exact test.

**Figure 2.**
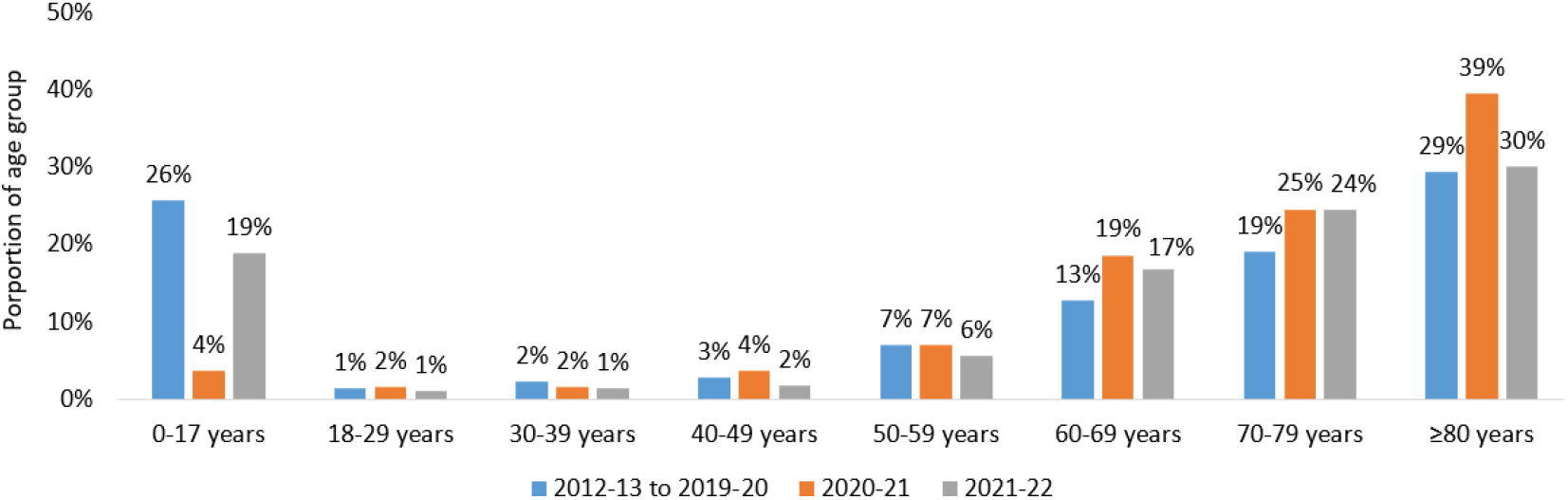
Age distribution of patients hospitalised for acute respiratory infections during pre-pandemic period (peaks of influenza seasons 2012-20) and two pandemic winter seasons (2020-21 and 2021-22) in Quebec, Canada

### Overall hospitalisations

During the pre-pandemic seasons, at least one respiratory virus was detected in 70.4% (4,107) patients **(Table 1**). The most frequently detected viruses were: influenza (2,107, 36.1%), RSV (1,080, 18.5%), hMPV (475, 8,1%), entero/rhinoviruses (460, 7.9%), and hCoV (376, 6.4%). Coinfections were detected in 720(12.3%) patients.

In 2020-21, significantly less patients (44.2%, 350) were positive for at least one virus (including SARS-CoV-2 and ORV) (**Table 1**). When considering only the weeks with LSPQ multiplex testing, 45.3% (279) were positive for at least one virus. The most frequently detected virus was SARS-CoV-2 (307, 38.8%). ORV were detected in 43 patients (5.4%), the most frequent were: entero/rhinoviruses (26, 3.3%), hMPV (11, 1.4%), and adenoviruses (10, 1.3%). No influenza and no RSV were detected. Coinfections were detected in 14 (1.8%) patients: 8 (1.0%) were a combination of ORV without SARS-CoV-2; 6 (0.8%) were SARS-CoV-2 with ORV.

In 2021-22, more patients (728, 62.2%) were positive for at least one virus, but the detection rate was still significantly lower than during the pre-pandemic (**Table 1**). The most frequently detected viruses were: SARS-CoV-2 (411, 35.1%), RSV (167, 14.3%), entero/rhinoviruses (106, 9.1%), adenoviruses (67, 5.7%), and bocaviruses (51, 4.4%). ORV (without SARS-CoV-2) were detected in 317 patients (27.1%). Coinfections were detected in 130 (11.1%) patients: 98 (8,4%) were a combination of ORV without SARS-CoV-2; 31 (2.7%) were SARS-CoV-2 with ORV. Similar to 2020-21, in 2021-22 virtually no influenza was detected up to epi-week 9, except for 1 influenza A(H3N2) virus in an adult patient (**Table 2A and 2B**). Hospitalisation due to RSV was detected much earlier (>10% during epi-weeks 40 to 48, with a maximum of 33% during epi-week 41) compared to pre-pandemic seasons (>10% during epi-weeks 52 to 10) (**Figure 1**).

**Table 2A.** Number of patients hospitalised for acute respiratory infections and detected viruses in Quebec, Canada, during pre-pandemic and pandemic winter seasons

| Number of patients and type of infection | Peaks of 2012-20 influenza seasons (4 hospitals) |  | 2020-21 (3 hospitals) | 2021-2022 (4 hospitals) |
| --- | --- | --- | --- | --- |
|  | Yearly average number (min-max) | Total number and detection rate, n(%) | Total number and detection rate, n(%) | Total number and detection rate, n(%) |
| Number of included patients | 186,6 (113 - 322) | 1493 (NA) | 29 (NA) | 221 (NA) |
| At least one respiratory virus* | 173 (103 - 307) | 1384 (92,7 %) | 17 (**58,6 %<sup>a</sup>**) | 201 (91,0 %) |
| Influenza, any | 45,5 (23 - 96) | 365 (24,4 %) | 0 (**0 %<sup>a</sup>**) | 0 (**0 %<sup>a</sup>**) |
| Influenza A | 34,3 (22 - 57) | 274 (18,4 %) | 0 (0 %) | 0 (**0 %<sup>a</sup>**) |
| H3N2 | 15,3 (0 - 32) | 122 (8,2 %) | 0 (0 %) | 0 (**0 %<sup>a</sup>**) |
| H1N1 | 18,5 (0 - 49) | 148 (9,9 %) | 0 (0 %) | 0 (**0 %<sup>a</sup>**) |
| A unsubtype | 0,5 (0 - 2) | 4 (0,3 %) | 0 (0 %) | 0 (0 %) |
| Influenza B | 11,4 (0 - 61) | 91 (6,1 %) | 0 (0 %) | 0 (**0 %<sup>a</sup>**) |
| RSV | 90,1 (44 - 172) | 721 (48,3 %) | 0 (**0 %<sup>a</sup>**) | 114 (51,6 %) |
| Adenovirus | 10,9 (4 - 20) | 87 (5,8 %) | 7 (**24,1 %<sup>a</sup>**) | 21 (9,5 %) |
| hMPV | 20,4 (0 - 38) | 163 (10,9 %) | 0 (0 %) | 16 (7,2 %) |
| hPIV 1-4 | 7,3 (0 - 13) | 58 (3,9 %) | 0 (0 %) | 14 (6,3 %) |
| hCoV | 19,3 (7 - 37) | 154 (10,3 %) | 3 (10,3 %) | 6 (**2,7 %<sup>a</sup>**) |
| Entero/rhinoviruses | 30 (22 - 48) | 240 (16,1 %) | 8 (27,6 %) | 70 (**31,7 %<sup>a</sup>**) |
| Bocaviruses | 16,9 (6 - 33) | 135 (9 %) | 6 (**20,7 %<sup>a</sup>**) | 45 (**20,4 %<sup>a</sup>**) |
| SARS-CoV-2 | NA | NA | 0 (0 %) | 18 (8,1 %) |
| Respiratory viruses without SARS-CoV-2 |  |  |  |  |
| Monoinfection | 116 (69 - 231) | 928 (62,2 %) | 10 (34,5 %) | 107 (48,4 %) |
| Co-infections, any RV without SARS-CoV-2 | 57 (27 - 91) | 456 (30,5 %) | 7 (24,1 %) | 76 (34,4 %) |
| SARS-CoV-2 |  |  |  |  |
| Monoinfection | NA | NA | 0 (0 %) | 10 (4,5 %) |
| Co-infections, SARS-CoV-2 + any RV | NA | NA | 0 (0 %) | 8<sup>b</sup> (3,6 %) |

\*Not mutually exclusive;
RSV: respiratory syncytial virus; hMPV: human metapneumovirus, hPIV: human parainfluenza viruses 1, 2, 3, and 4, hCoV: common human coronaviruses; RV: respiratory viruses;
<sup>a</sup> p value <0.05 for comparison with 2012-20, Fisher exact test;
<sup>b</sup> SARS-CoV-2 in coinfection with: adenoviruses n=4, HMPV n=2, Entero/rhinoviruses n=2.

B Adults
| Number of patients and type of infection | Peaks of 2012-20 influenza seasons (4 hospitals) |  | 2020-21 (3 hospitals) | 2021-2022 (4 hospitals) |
| --- | --- | --- | --- | --- |
|  | Yearly average number (min-max) | Total number and detection rate, n(%) | Total number and detection rate, n(%) | Yearly average number (min-max) |
| Number of included patients | 542,4 (388 - 689) | 4339(NA) | 762 (NA) | 949 (NA) |
| At least one respiratory virus* | 340,4 (224 - 425) | 2723 (62,8 %) | 333 (**43,7 %<sup>a</sup>**) | 527 (**55,5 %<sup>a</sup>**) |
| Influenza, any | 217,1 (132 - 334) | 1742 (40,1 %) | 0 (**0 %<sup>a</sup>**) | 1 (**0,1 %<sup>a</sup>**) |
| Influenza A | 185,4 (109 - 324) | 1483 (34,2 %) | 0 (**0 %<sup>a</sup>**) | 1 (**0,1 %<sup>a</sup>**) |
| H3N2 | 125,8 (2 - 316) | 1006 (23,2 %) | 0 (**0 %<sup>a</sup>**) | 1 (**0,1 %<sup>a</sup>**) |
| H1N1 | 57 (0 - 123) | 456 (10,5 %) | 0 (**0 %<sup>a</sup>**) | 0 (**0 %<sup>a</sup>**) |
| A unsubtype | 2,6 (0 - 8) | 21 (0,5 %) | 0 (0 %) | 0 (**0 %<sup>a</sup>**) |
| Influenza B | 32,4 (1 - 146) | 259 (6,0 %) | 0 (0 %<sup>a</sup>) | 0 (**0 %<sup>a</sup>**) |
| RSV | 44,9 (19 - 75) | 359 (8,3 %) | 0 (0 %<sup>a</sup>) | 53 (**5,6 %<sup>a</sup>**) |
| Adenovirus | 1,6 (0 - 5) | 13 (0,3 %) | 3 (0,4 %) | 46 (**4,9 %<sup>a</sup>**) |
| hMPV | 39 (2 - 103) | 312 (7,2 %) | 11 (**1,4 %<sup>a</sup>**) | 14 (**1,5 %<sup>a</sup>**) |
| hPIV 1-4 | 14,5 (3-25) | 116 (2,7 %) | 0 (**0 %<sup>a</sup>**) | 11 (**1,2 %<sup>a</sup>**) |
| hCoV | 27,8 (7 - 49) | 222 (5,1 %) | 0 (**0 %<sup>a</sup>**) | 13 (**1,4 %<sup>a</sup>**) |
| Enteroviruses | 27,5 (12 - 43) | 220 (5,1 %) | 18 (**2,4 %<sup>a</sup>**) | 36 (3,8 %) |
| Bocaviruses | 2,5 (0 - 6) | 20 (0,5 %) | 2 (0,3 %) | 6 (0,6 %) |
| SARS-CoV-2 | NA | NA | 307 (40,3 %) | 393 (41,4 %) |
| Respiratory viruses without SARS-CoV-2 |  |  |  |  |
| Monoinfection | 307,4 (205) | 2459 (56,7 %) | 25 (3,2 %) | 112 (11,8 %) |
| Co-infections, any RV without SARS-CoV-2 | 33 (12) | 264 (6,1 %) | 1 (0,1 %) | 22 (2,3 %) |
| SARS-CoV-2 |  |  |  |  |
| Monoinfection | NA | NA | 301 (39,5 %) | 370 (39,0 %) |
| Co-infections, SARS-CoV-2 + any RV | NA | NA | 6<sup>b</sup> (0,8 %) | 23<sup>c</sup> (2,4 %) |

\*Not mutually exclusive;
RSV: respiratory syncytial virus; hMPV: human metapneumovirus, hPIV: human parainfluenza viruses 1, 2, 3, and 4; hCoV: common human coronaviruses; RV: respiratory viruses;
<sup>a</sup> p value <0.05 for comparison with 2012-20, Fisher exact test;
<sup>b</sup> SARS-CoV-2 in coinfection with: hMPV n=4, adenoviruses n=2, enteroviruses n=1, bocaviruses n=1;
<sup>c</sup> SARS-CoV-2 in coinfection with: adenoviruses n=15, hPIV n=3, hCoV n=2, enteroviruses n=2, RSV n=1.

### Pediatric hospitalisations

While during pre-pandemic period at least one respiratory virus was detected in 92.7% (1,384) children including 30.5% (456s) coinfections, detection rate decreased to 58.6% (29) in 2020-21 (24.1% coinfections), and increased again to 91.0% (221) in 2021-22 (38.0% coinfections).

Prior to COVID-19 pandemic, the most frequently detected viruses in hospitalised children were RSV (48.3%) and influenza (24.4%) (Table 2A). In 2020-21 all hospitalisations with an identified virus in children were due to respiratory viruses other than SARS-CoV-2, mostly entero/rhinoviruses (27.6%,), adenoviruses (24.1%), and bocaviruses (20.7%). Proportions of detected adenoviruses and bocaviruses were significantly higher than in previous seasons (**Table 2A**); no significant differences were detected for the rest of viruses. In 2021-22 most hospitalisations in children were due to ORV without SARS-CoV-2 (83%(183)); only 8%(18) were for SARS-CoV-2 (including 8 coinfections with ORV). The predominant virus was RSV (51.6%) followed by entero/rhinoviruses (31.7%) and bocavirus (20.4%). Compared to pre-pandemic seasons, significantly more entero/rhinoviruses and bocaviruses and less hCoV were detected (**Table 2A**).

### Adult hospitalisations

During the pre-pandemic seasons, at least one respiratory virus was detected in 63% (2,723) adults (6.1% coinfections); influenza was by far the most predominant virus (40.1%).

In 2020-21, significantly less adults (44% (333)) were positive for at least one virus (including SARS-CoV-2 and ORV) (0.9% coinfections). Proportions of at least one virus detected were significantly lower in 18-29-year-olds and ≥60-year-olds, but similar in 30-59-year-olds (**Table 1**). The SARS-CoV-2 was the main cause of ARI hospitalization in adults (40.3%(307)) and the only cause of hospitalisation in 30-49-year-olds (**Table 1 and 2**). At least one ORV with or without SARS-CoV-2 was detected in 4.2% (32) adults, most frequently entero/rhinoviruses (2.4%) and hMPV (1.4%). Significantly lower proportions of hMPV, hPIV, hCoV and entero/rhinoviruses compared to pre-pandemic period were detected (**Table 2B**). In 2021-22, significantly less adults (55.5%, 527) were positive for at least one virus (4.7% coinfections) compared to pre-pandemic period; proportions were similar in 18-59-year-olds and ≥80-year-olds, and significantly lower in 60-79-year-olds.

Similar to the 1^st^ pandemic season, the SARS-CoV-2 was the most frequent virus in adults (41.4%, 393). ORV with or without SARS-CoV-2 were detected in all examined adult age groups except the 40-49-year-olds, for an overall proportion of 16.5% (157) (**Table 1**). Compared to the 1^st^ pandemic season, the SARS-CoV-2 was detected more often in adults younger than 50 years (the only significant difference in 18-29-year-olds, **Table 1**); detection was similar in adults 50 years and older. Compared to pre-pandemic period, significantly more adenoviruses, and less hMPV, hPIV1-4, and hCoV were detected (**Table 2B**).

Results of viral detection in 2 additional hospitals in 2021-22 compared to 4 hospitals included in the main analysis are presented in **Supplementary table 2**. Differences might be explained by different assays, populations (only pediatric in one hospital, only adult in the other hospital) and timespan. While comparisons with previous seasons are not appropriate in this context, we included results from additional hospitals in the presentation of viral etiology by week in order to illustrate a more comprehensive impact of RV on hospitalisations (**Figure 1**).

## Discussion

Our report, based on systematic RV detection, describes changes in the etiology (17 RV and SARS-CoV-2) of pediatric and adult ARI hospitalisations during 2 pandemic years compared with 8 pre-pandemic years in the same population. To our knowledge, this is the longest time span of follow-up during both pandemic and pre-pandemic periods in both pediatric and adult hospitalised patients. We detected significant shifts in age distribution and continuing changes in viral etiology during the two pandemic years compared to pre-pandemic period, but also between the first and the second pandemic year. While overall the SARS-CoV-2 was the most frequent viral etiology of ARI hospitalisations during both pandemic years, it was absent in children during the first year but present during the second year. Detected differences in age distribution and etiology of respiratory hospitalisations are a consequence of changes in both ORV and SARS-CoV-2 circulation, reflecting the intensity of mitigation measures. For instance, the circulation of ORV was very low in 2020-21 when more stringent measures were implemented, it increased during the autumn 2021 following the easing of some measures, declined in December 2021 following the tightening of the measures in response to the omicron wave and increased again in February-March 2022 following broader travelling and school opening (**Supplementary Table 1**).

Another potential contributive factor was the SARS-CoV-2 variant evolution. For example, ancestral and alpha SARS-CoV-2 variants circulated during the 2020-21 season and affected mostly adults; while delta, and then omicron variants, both affecting more importantly pediatric population, predominated during the autumn and the winter of 2021-22(24). Also, an unhabitual surge of RSV in August-September 2021(27) resulted in higher proportion of children hospitalised with both ORV (predominantly RSV) and COVID-19 compared to the first pandemic season. Finally, COVID-19 vaccines and outpatient antiviral availability and uptake, effective to prevent hospitalisations, varied by age group. During the second pandemic year, older patients were prioritized in COVID-19 vaccination campaigns and more patients with comorbidities were offered outpatient antiviral treatment. For children, however, COVID-19 vaccines were available later and in young adults vaccine uptake was lower than in older population (28).

Overall during the pre-pandemic influenza seasons, the two most frequently detected viruses among patients hospitalised with ARI were influenza and RSV. The SARS-CoV-2 was the most important respiratory virus during the first pandemic year; in 2021-22 its contribution decreased while contribution of ORV increased. However, there were differences in pediatric and adult population. RV (mostly RSV) affected more children than adults during pre-pandemic winter seasons, and although their overall impact was lower during pandemic period, children remained mostly affected by ORV and not by the SARS-CoV-2 during both pandemic years. In adults, influenza was the most important virus during the pre-pandemic years, while SARS-CoV-2 was by far more important than ORV during the two pandemic years.

The important reduction of pediatric respiratory hospitalisations, virtual absence of influenza and RSV during first pandemic year and increased role of COVID-19 and ORV, especially RSV, in hospitalised children during second pandemic year in our study are in line with reports from some other countries and other provinces of Canada (13,16,17,29,30). Of note, the timing of the unusual surge of RSV varied by region of the world: in Europe, Israel and USA increases started to be reported in the Spring-Summer of 2021(13,16,17,29). Major shifts in the epidemiology of RSV with large-scale outbreaks were reported in New Zealand (corresponding to usual season)(31) and Australia (out-of-season)(32). In Canada, the first province to report a surge of RSV was Quebec in August 2021, while the other provinces followed with a delay of 2-3 months(30).

This study has limitations. First, the pre-pandemic surveillance occurred during the peaks of influenza activity and therefore relative contribution of other RV may be underestimated as compared to entire winter seasons. Surveillance periods during pandemic years were mostly tailored to the increase in respiratory hospitalisations following increased SARS-CoV-2 circulation and were much longer. However, we think the comparison is still valid because it includes periods with most strain on hospital capacity due to intensive circulation of either ORV, SARS-CoV-2, or both. Second, ARI definition used during pandemic years was broader than during pre-pandemic years and may have contributed to lower detection of RV. Third, the first pandemic year was limited to only 3 hospitals and periodic pauses and sampling of the enrolment during the 2 pandemic years were necessary given the stretch of resources which limited the sample size. However, this should not have influenced relative contribution of RV and global comparisons as the surveillance period during pandemic years was much longer than previous seasons. Also, this approach allowed the maintenance of the surveillance in the context of resource challenges during the pandemic. Finally, our results may not necessarily be extrapolated to other regions or periods of time because of temporal and geographical differences in ORV and SARS-CoV-2 epidemiology. For example, we do not present here the last weeks of the 2021-22 when a late-season influenza outbreak occurred in Quebec (27). The main strength of this study was the systematic testing for a broad panel of viruses for all admitted patients with symptoms of ARI during the surveillance periods. Moreover, it included both pediatric and adult population during a total of 10 years, with the participation of at least 3 hospitals from different Quebec regions in each year, and the possibility to distinguish community-acquired and nosocomial infections.

In conclusion, important shifts in age distribution and viral etiology of ARI hospitalisations in children and adults were observed during the two first pandemic years. While the first pandemic year was significantly different from the pre-pandemic winter seasons, the second year was more comparable, both in terms of age distribution and RV contribution. The complex interplay between mitigation measures, intrinsic seasonality and secular trends of ORV, changes in circulating SARS-CoV-2 variants and their severity, COVID-19 vaccine uptake and effectiveness, outpatient antiviral treatments and potential viral interference (33,34) are all factors that may contribute to variations of ORV and SARS-CoV-2 role in hospital morbidity. Even if we do not report here the results for the entire 2021-22 season, our study underscores importance of surveillance in understanding altered seasonal patterns of RV and shows that the role of SARS-CoV-2 relative to ORV is continually changing. Current situation may reflect a transition period until it will find its ecological niche in human population. With the continued easing of mitigation measures, an increasing role of usual winter viruses is expected. Although new SARS-CoV-2 variants may emerge and cause occasional increases in hospitalisations, in the long run it may establish itself as another respiratory virus. At this point, it is difficult to foresee its role compared to ORV. Increased scrutiny to continuing changes in the etiology of ARI hospitalisations is needed in order to inform mathematical modelling and appropriate public health recommendations.

## Data Availability

All data produced in the present study are available upon reasonable request to the authors conditional on the approval of Ministry of Health of Quebec

## Acknowledgments

We acknowledge France Bouchard who coordinated the surveillance and Sophie Auger who entered and cleaned collected data throughout all years. We are extremely grateful to the front-line surveillance staff from the participating hospitals who collected and provided data in challenging circumstances during the pandemic period: Ménard Francois, Gagnon Maude, Desbiens Karine, Tremblay Chantale, Simard Patricia, Murry Carole, Rhainds Jennifer (Chicoutimi hospital); Estel Deblois, Alexandra Bouffard, Alexandra Fortier, Alexandra Mondor, Rosalie Beaudoin, Kristina Boucher, Mélyna Carrier, Brigitte Dion, Marie-Ève Chamberland, Lise Anne Paradis (Hôtel-Dieu de Lévis hospital); Dolcé Patrick, Gagnon Isabelle, Lévesques Julie, Bernatchez Isabelle, Lévesques Janie (Rimouski hospital); Poirier André, Danylo Alexis, Tapps Danielle, Loranger Josée, Toupin Guylaine (Trois-Rivières hospital); Wadas Katerin, Lafleur Caroline, Ane Tres Silicia, Tomas Fernanda (CUSM); Thibeault Roseline, Jacob-Wagner Marieve, Hamelin Marie-Eve, Theriault Ariane, Pelletier-Bélanger Joannie, Côté Claudia (CHUL). We thank also Desautels Lyne, Martineau Christine, Ménard Joël from LSPQ who supported the surveillance despite the exponentially increasing demands on laboratories during the pandemic. We also acknowledge the support of Hany Geagea, Lauriane Padet, and Radhouene Doggui for revising the literature.

