## Supplementary data for "The changing landscape of respiratory viruses contributing to respiratory hospitalisations: results from a hospital-based surveillance in Quebec, Canada, 2012-13 to 2021-22"

**Supplementary Figure 1** Number patients hospitalised for acute respiratory infections included in the surveillance in Quebec, Canada, by epi-week during the pre-pandemic (A) and pandemic (B) periods

**A**

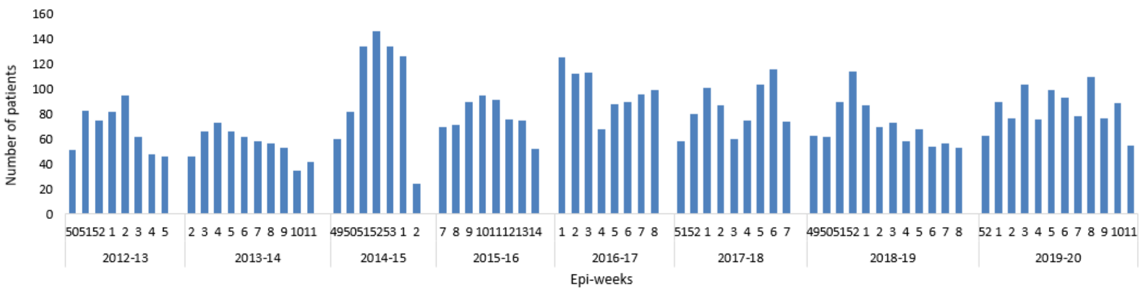

**B**

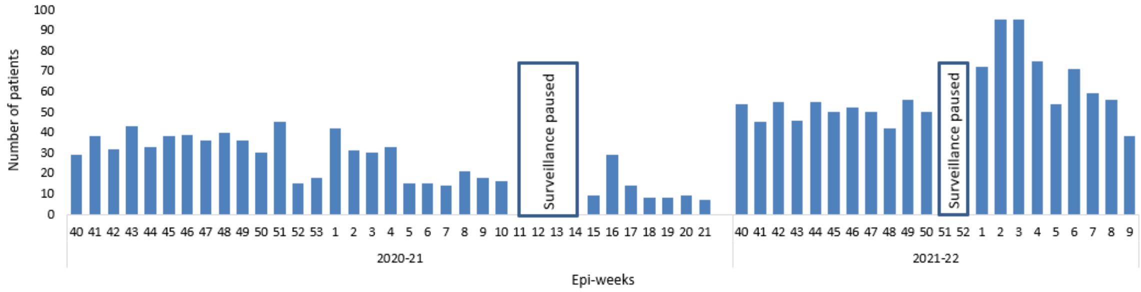

Note: Four hospitals participated during pre-pandemic period. Three hospitals participated during 2020-21 with periodic sampling by day of week. Four hospitals participated during 2021-22 with periodic sampling by day of week. The two additional hospitals are not presented.

**Supplementary Figure 2** Surveillance flowchart by period*

| Period | 2012-2020 (pre-pandemic) | 2020-2021 (first pandemic year) | 2021-2022 (second pandemic year) |
|---|---|---|---|
| **Number of swabbed patients** | 6,412 | 1,454 | 1,521 |
| **Exclusions:** | | | |
| Refusal/unable to consent | 68 | NA | NA |
| Eligibility criteria not met[1] | 51 | 63 | 182 |
| Enrolled before the surveillance period | 0 | 26 | 52 |
| Missed by nurses | 25 | NA | NA |
| Admitted less than 24 hours | 18 | 2 | 16 |
| COVID-19 patient transferred from another region[2] | NA | 222 | NA |
| Health-care acquired ARI[3] | 364 | 145 | 54 |
| Samples not received by LSPQ/insufficient volume | 36 | 205 | 22 |
| Other reason[4] | 18 | 0 | 25 |
| **Included in the analysis** | 5,832 | 791 | 1,170 |

*4 centers included

NA: not applicable

[1]For example: symptoms related to another condition (i.e fever in urinary tract infection, cellulitis)

[2]Transfers from centers not designated to receive COVID-19 patients according to government mandate situated in regions not covered by participating hospitals outside the COVID-19 mandate

[3]Defined as: Onset of ARI symptoms >72 hours after admission

[4]For example: duplicate cases, medical chart not available.

**Supplementary table 1 Timeline of SARS-CoV-2 epidemiology and main mitigation measures in Quebec** (more details available at https://www.inspq.qc.ca/covid-19/donnees/ligne-du-temps).

| Year | Date | Events |
|---|---|---|
| 2020 | **February 23** | **Start of first wave** |
| | March 13 | Declaration of health emergency by the province of Quebec |
| | March 14 | Cancellation of non-essential visits to long-term-care-facilities (LTCF) and hospitals; stay-at-home order for ≥70 years, physical distancing; closure of some public spaces, school and kindergartens closure |
| | March 18 | International border closure |
| | March 18 | LTCF and nursing homes lockdown, progressive closure of all services excepting those essential; restrictions of travel between some regions with check points at entry; cancellation of all summer festivals and activities |
| | April 15 | Progressive opening of some sectors, easing of some measures |
| | **July 11** | **End of first wave** |
| | July 13 | Mandatory use of masks or face covering in public transport |
| | July 18 | Mandatory use of masks or face covering in all public enclosed spaces |
| | **August 20** | **Start of second wave** |
| | September 8 | Graded regional warning system in place |
| | September 11 | Progressive tightening of measures |
| | December 14 | Start of COVID-19 vaccination with LTCF residents and LTCF health-care workers (HCW) according to priority order |
| | December 25 | Closure of non-essential businesses |
| | December 29 | First detection of the B.1.1.7 variant (Alpha) |
| | December 31 | Given limited number of COVID-19 vaccines, dose 1 is prioritized in order to rapidly achieve higher coverage in vulnerable groups |
| 2021 | February 8 | Progressive easing of measures |
| | February 9 | First detection of the B.1.351 variant (Beta) |
| | March 1 | Vaccination extended to general population according to priority groups |
| | **March 20** | **End of second wave** |
| | **March 21** | **Start of third wave** |
| | March 25 | Only one dose of vaccine may be administered to persons with prior COVID-19 infection |
| | April 1 | Special emergency measures in some regions |
| | April 26 | First detection of the B.1.617 variant (Delta) |
| | May 18 | 50% of Quebec population ≥12 years received at least 1 dose |
| | June 6 | 75% of Quebec population ≥12 years received at least 1 dose |
| | **July 17** | **End of third wave** |
| | **July 18** | **Start of forth wave** |
| | September 1 | Vaccination passport required for access to most public spaces |
| | September 28 | Booster dose recommended for LTCF and nursing homes residents |
| | September 30 | 75% of Quebec population is adequately vaccinated |
| | November 1 | 6 months between booster dose and last dose received is recommended. Optimal interval between 1$^{st}$ and 2$^{nd}$ dose is 8 weeks |
| | November 15 | Progressive easing of some measures |

| | | |
|---|---|---|
| | November 16 | Booster dose recommended to ≥70 years from community, persons who received a viral vector vaccine may receive a mRNA vaccine booster |
| | November 24 | Start of 5-11-year-olds vaccination |
| | November 29 | Detection of omicron variant |
| | December 4 | **End of forth wave** |
| | December 5 | **Start of fifth wave** |
| | December 20 | Progressive tightening of measures, including curfew starting December 31<br>Booster dose advanced for all ≥ 60 years; interval between last dose and booster shortened from 6 to 3 months |
| | December 29 | Booster dose offered to essential workers followed-up by all population according to age groups |
| 2022 | January 5 | Change in PCR screening priorities and increased access to self-testing |
| | January 17 | Curfew lifted |
| | January 31 | Progressive easing of some of the measures |
| | February 16 | Vaccination passports no longer required for some public spaces |
| | February 18 | Booster dose offered to 12-17-year-olds (at least 3 months from the last dose) |
| | February 21-March 7 | Further easing of measures |

**Supplementary table 2** Results of viral detection in patients hospitalised for acute respiratory infections in 2021-2022, 4 hospitals participating during pre-pandemic period and 2 additional hospitals

| Number of patients and type of infection | 2021-22 (4 main hospitals) | 2021-22 (2 additional hospitals) |
|---|---|---|
| | Total number and detection rate, n(%) | Total number and detection rate, n(%) |
| Number of tested patients | 1,170 | 594 |
| At least one respiratory virus | 728(62.2%) | 446(75.1%)* |
| Influenza, any | 1(0.1%) | 1(0.2%) |
| RSV | 107(9.1%) | 27(4.5%)* |
| Adenovirus | 19(1.6%) | 12(2.0%) |
| hMPV | 15(1.3%) | 33(5.6%)* |
| hPIV 1-4 | 7(0.6%) | 7(1.2%) |
| hCoV | 12(1.0%) | 8(1.3%) |
| Entero/rhinoviruses | 49(4.2%) | 84(14.1%)* |
| Bocaviruses | 9(0.8%) | 2(0.3%) |
| SARS-CoV-2 | 380(32.5%) | 196(33.0%) |
| Respiratory viruses without SARS-CoV-2 | | |
| Monoinfection | 599(51.2%) | 373(62.8%)* |
| Co-infections, any RV without SARS-CoV-2 | 98(8.4%) | 55(9.3%) |
| SARS-CoV-2 | | |
| Monoinfection | 349(29.8%) | 178(30.0%) |
| Co-infections, SARS-CoV-2 + any RV | 31(2.6%) | 18(3.0%) |

RSV: respiratory syncytial virus; hMPV: human metapneumovirus, hPIV: human parainfluenza viruses 1, 2, 3, and 4, hCoV: common human coronaviruses; RV: respiratory viruses;
*P<0,0001
